# A method for complete characterization of complex germline rearrangements from long DNA reads

**DOI:** 10.1101/19006379

**Authors:** Satomi Mitsuhashi, Sachiko Ohori, Kazutaka Katoh, Martin C Frith, Naomichi Matsumoto

## Abstract

Many genetic/genomic disorders are caused by genomic rearrangements. Standard methods can often characterize these variations only partly, e.g. copy number changes. We describe full characterization of complex chromosomal rearrangements, based on whole-genome-coverage sequencing of long DNA reads from four patients with chromosomal translocations. We developed a new analysis pipeline, which filters out rearrangements seen in humans without the same disease, reducing the number of loci per patient from a few thousand to a few dozen. For one patient with two reciprocal chromosomal translocations, we find that the translocation points have complex rearrangements of multiple DNA fragments involving 5 chromosomes, which we could order and orient by an automatic algorithm, thereby fully reconstructing the rearrangement. Some important properties of these rearrangements, such as sequence loss, are holistic: they cannot be inferred from any part of the rearrangement, but only from the fully-reconstructed rearrangement. In this patient, the rearrangements were evidently caused by shattering of the chromosomes into multiple fragments, which rejoined in a different order and orientation with loss of some fragments. Our approach promises to fully characterize many congenital germline rearrangements, provided they do not involve poorly-understood loci such as centromeric repeats.

## Introduction

Various germline DNA sequence changes are known to cause rare genetic disorders. Many small nucleotide-level changes (one to a few bases) in 4,143 genes have been reported in OMIM (https://www.omim.org/) (as of Aug 24, 2019), which are known as single gene disorders. In addition to these small changes, large structural variations of the chromosomes can also cause diseases.

Previous studies on pathogenic structural changes in patients with genetic/genomic disorders found chromosomal abnormalities by microscopy, by detecting copy number variations (CNVs) using microarrays^1^, or by detecting both CNVs and breakpoints using high-throughput short read sequencing^2^. However, there are difficulties in precisely identifying sequence-level changes especially in highly similar repetitive sequences (e.g. simple repeats, recently-integrated transposable elements), or in finding how these rearrangements are ordered^3^. Long read sequencing (PacBio or nanopore) is advantageous for characterizing rearrangements in such cases, and is recently beginning to be used for patient genome analysis to identify pathogenic variations^4-6^. In addition, if rearrangements are complex (e.g. chromothripsis), long read sequencing (reads are more than 10 kb in length) has a further advantage because one read may encompass all or much of a complex rearrangement^7^. Chromothripsis is a chaotic complex rearrangement, where many fragments of the genome are rearranged into derivative chromosomes. Current approaches to analyze chromothripsis usually need manual inspection to reconstruct whole rearrangements. Detection and reconstruction methods for complex rearrangements are needed to characterize pathogenic variations from whole genome sequencing data.

In order to understand rearrangements between two sequences (e.g. a read and a genome), we must determine equivalent positions, i.e. bases descended from the same base in the most recent common ancestor of the sequences. This is not necessarily easy, due to sequences that are similar but not equivalent (e.g. alpha-1 and alpha-2 globin). If we compare two sequences that have both undergone deletions, duplications, and rearrangements since their common ancestor, it seems hard to reliably determine equivalent bases. To make the problem tractable, we impose an assumption: that we are comparing a derived sequence (a DNA read) to an ancestral sequence (the genome)^8^. This means that every part of the read is descended from (equivalent to) a unique part of the genome. (The exception is “spontaneously generated” sequence not descended from an ancestor: this is rare, and we allow for it by allowing parts of the read to not align anywhere.) Thus, we need to accurately: divide the read into (one or more) parts and align each part to the genome. To do this, we first learn the rates of small insertions, deletions, and each kind of substitution in the reads^9^, then find the most-likely division and alignment based on these rates^8,10^. We can also calculate the probability that each base is wrongly aligned, which is high when part of a read aligns almost equally well to several genome loci. This approach was previously used to characterize rearrangements that are “localized”, i.e. encompassed by one DNA read^8^.

Here we extend this approach, to: find arbitrary (non-localized) rearrangements, subtract rearrangements found in control individuals, then order and orient rearranged DNA reads to fully reconstruct complex rearrangements in derivative chromosomes.

## Results

### Nanopore sequencing of 4 patients with chromosomal translocations

We sequenced genomic DNA from 4 patients with reciprocal chromosomal translocations using a nanopore long read sequencer, PromethION (Supplementary Table 1). Clinical information of these patients is described in the Supplementary methods and elsewhere^11-14^. We applied newly developed software, `dnarrange` (https://github.com/mcfrith/dnarrange) to find and characterize DNA sequence rearrangements in these patients. `dnarrange` finds DNA reads that have rearrangements relative to a reference genome, and groups reads that overlap the same rearrangement (Supplementary Methods). Then, it filters out rearrangements that are seen in any of 33 control individuals (Fig1, Supplementary Table 1). The number of read groups decreased exponentially with the first several controls, then stabilized, which suggests that there are numerous commonly shared rearrangements in the population (Fig2b, Fig3b, Fig4b, Fig5b, Supplementary Table 2). Next, we merged (a.k.a. assembled) the reads of each group into a consensus sequence using `lamassemble` (https://gitlab.com/mcfrith/lamassemble), and realigned to the reference genome. Representative examples of detected rearrangements are shown with raw reads and consensus sequences in Supplementary Fig 1.

**Figure 1.**
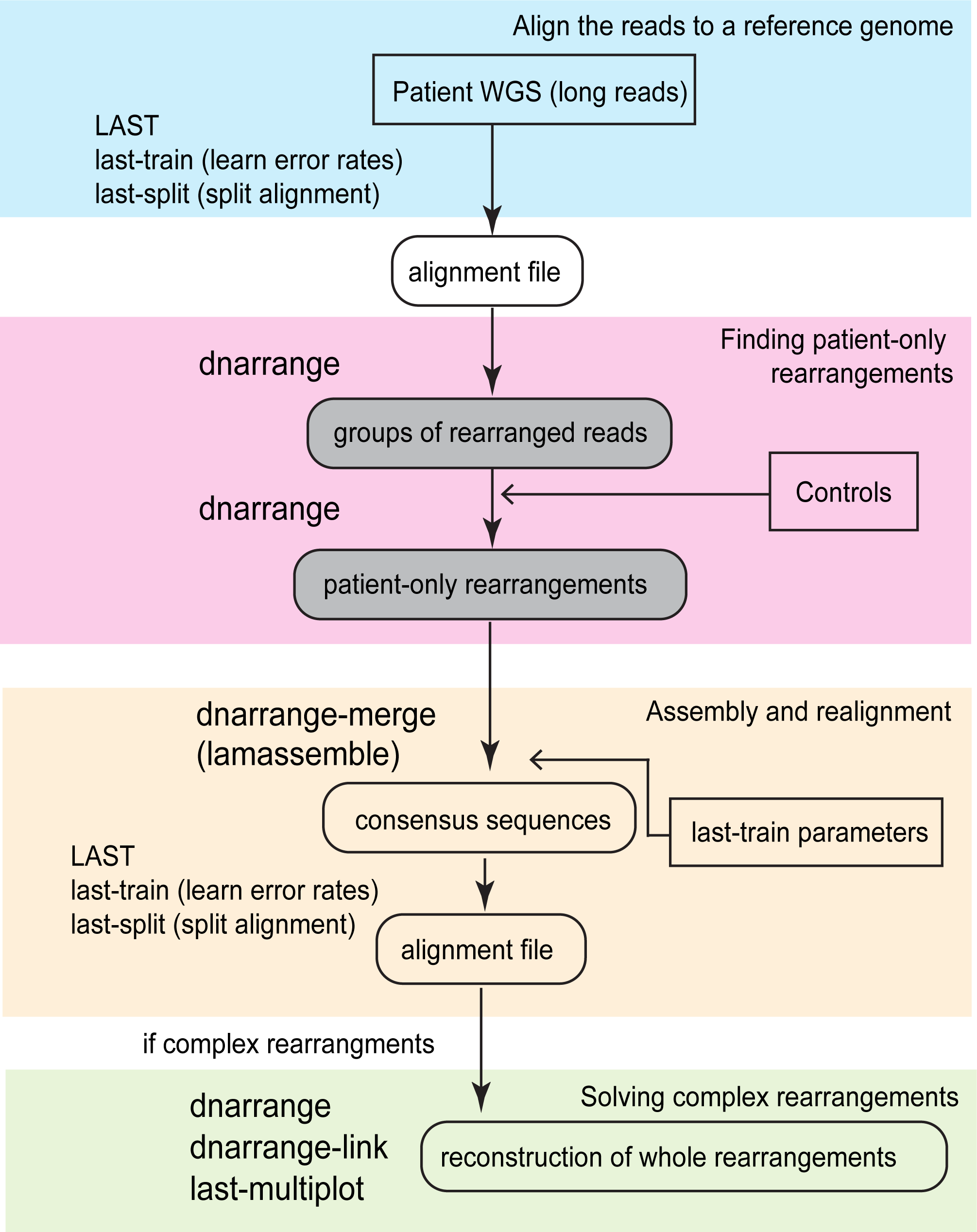
Schematic diagram of chromosomal rearrangement analysis pipeline. Long DNA reads are aligned to a reference genome using LAST (blue box), then `dnarrange` finds rearranged reads, and groups reads that overlap the same rearrangement (pink box). `lamassemble` merges/assembles each group of reads into a consensus sequence (yellow box). When there is a “complex” rearrangement (more than one group of rearranged reads is needed to understand the full structure of the rearrangement), `dnarrange-link` was used to infer the order and orientation of the groups, and thereby reconstruct derivative chromosomes (green box).

**Figure 2.**
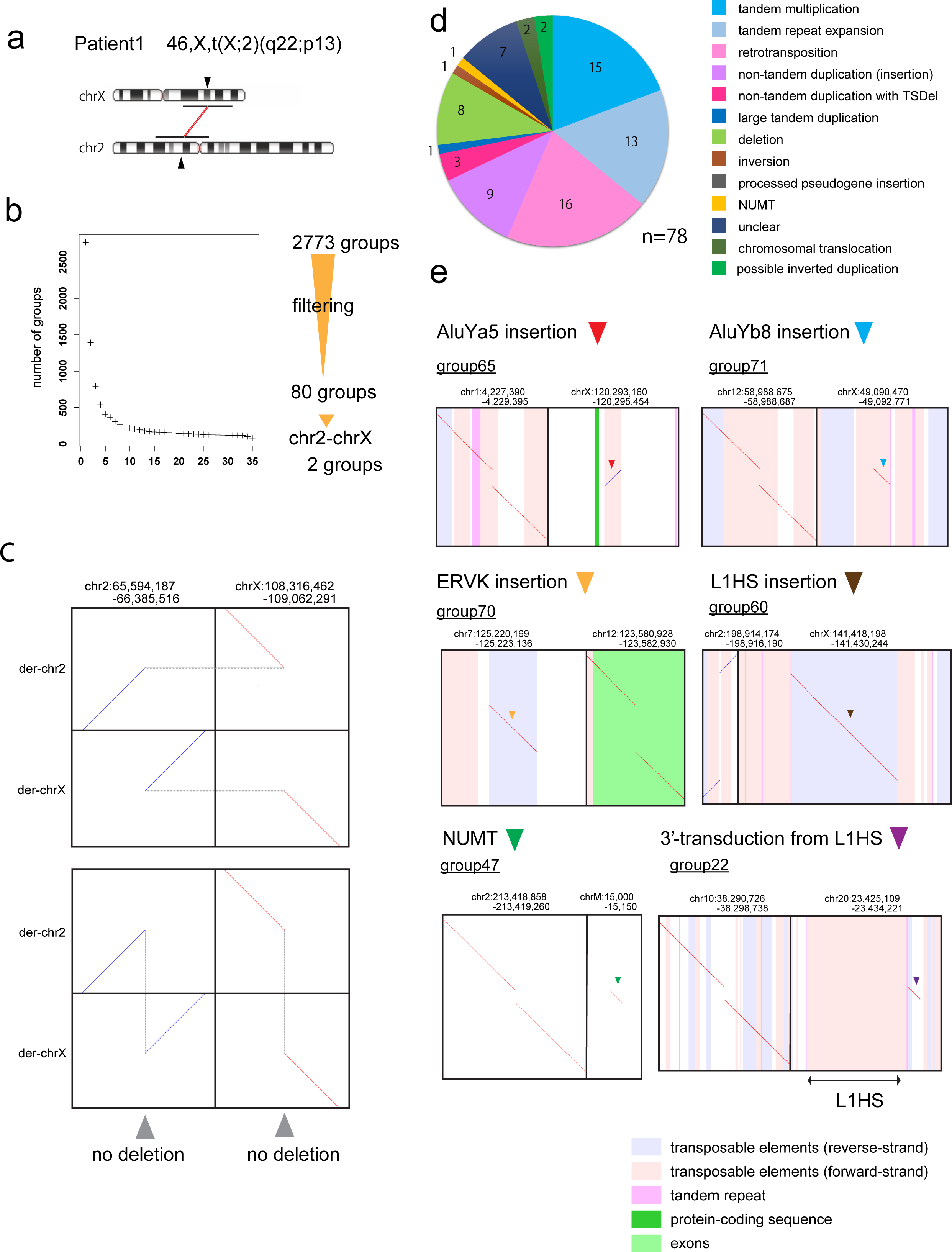
Chromosomal rearrangement in Patient 1 with 46,X,t(X;2)(q22;p13) a. Ideograms showing Patient 1’s translocation between chrXq22 and chr2p13. Chromosome images are from NCBI genome decoration (https://www.ncbi.nlm.nih.gov/genome/tools/gdp). b. Filtering out rearrangements shared with 33 controls. Finally, 80 groups of reads with patient-only rearrangements are found. Two of the 80 groups show reciprocal chr2-chrX translocation. c. Dot-plot of reconstructed derivative chromosomes shows reciprocal balanced chromosomal translocation. (Upper panel: horizontal dotted gray lines join the parts of each derivative chromosome. Lower panel: vertical dotted gray lines join fragments that come from adjacent parts of the reference genome, showing there is no large deletion or duplication). d. Pie chart of the types of rearrangement. TSDel: target site deletion. NUMT: nuclear mitochondrial DNA insertion. e. Examples of retrotransposition and NUMT insertion. (The alignments to retrotransposons, e.g. the AluYa5 in chrX, often have low confidence, indicating uncertainty that this specific AluYa5 is the source.)

**Figure 3.**
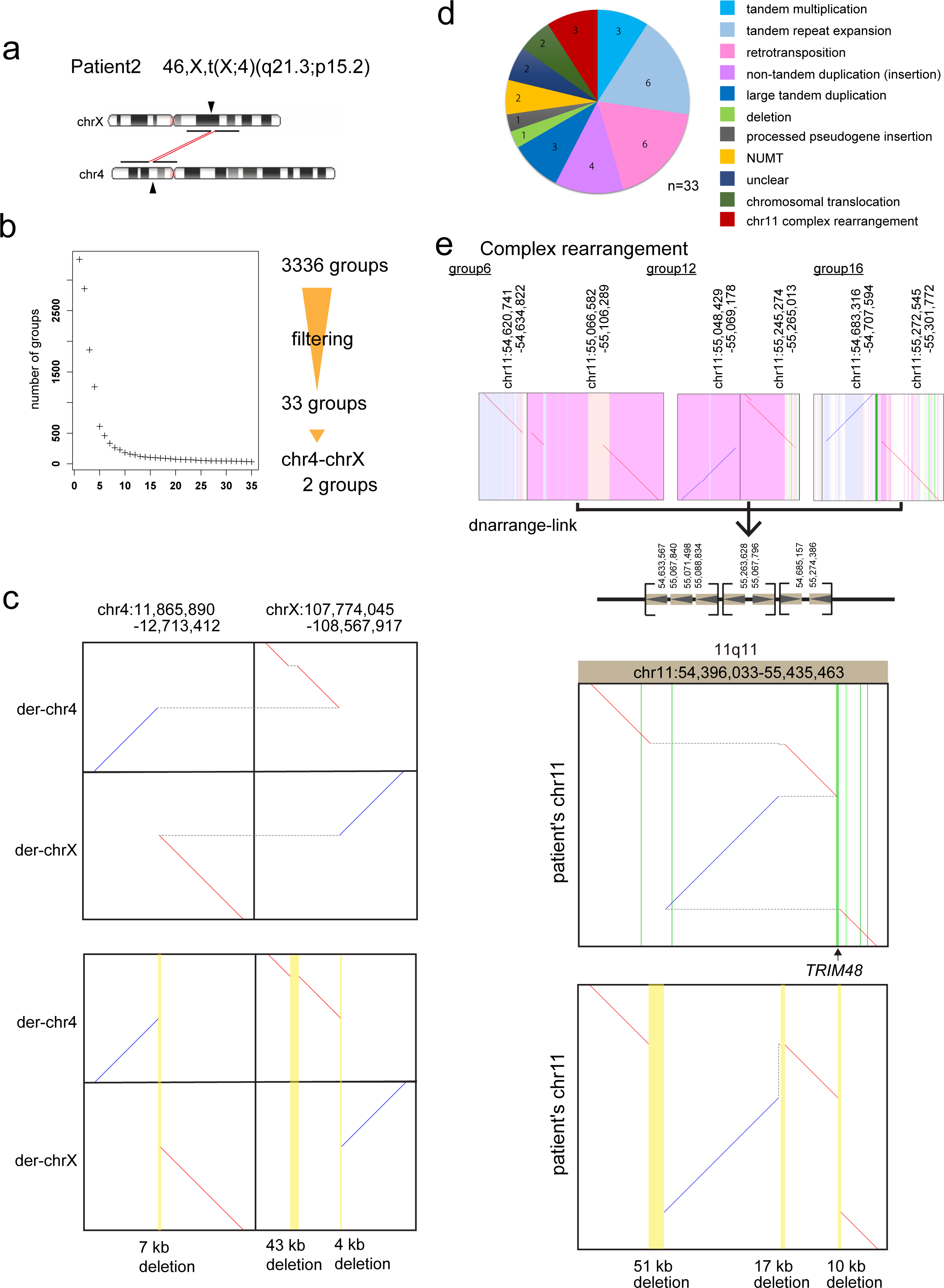
Chromosomal rearrangement in Patient 2 with 46,X,t(X;4)(q21.3;p15.2) a. Ideograms showing Patient 2’s translocation between chr4p.15.2 and chrXq21.3. b. Filtering out rearrangements shared with controls produces 33 groups of reads with patient-only rearrangements. Two of the 33 groups show chr4-chrX translocation. c. Dot-plot of derivative (vertical) versus ancestral/reference (horizontal) chromosomes showing reciprocal chromosomal translocation. There are 7 kb and 4 kb deletions at the breakpoints in chr4 and chrX, respectively. There is also a 43 kb deletion in chrX. Yellow vertical lines show deletions. d. Pie chart of patient-only rearrangements. e. A complex rearrangement on chr11. Three dotplots at chr11q11 were linked to reconstruct a complex rearrangement with three sequence losses, chr11:54633567-54685157 (51 Kb), chr11:55071498-55088834 (17 Kb) and chr11:55263629-55274386 (10 Kb). The latter disrupts the *TRIM48* gene, which is not known to cause any diseases. Upper dot-plot panel: horizontal dotted gray lines join the parts of each derivative chromosome. Green vertical lines show exons. Lower dot-plot panel: vertical dotted gray lines join fragments that come from adjacent parts of the reference genome. Yellow vertical lines show deletions.

**Figure 4.**
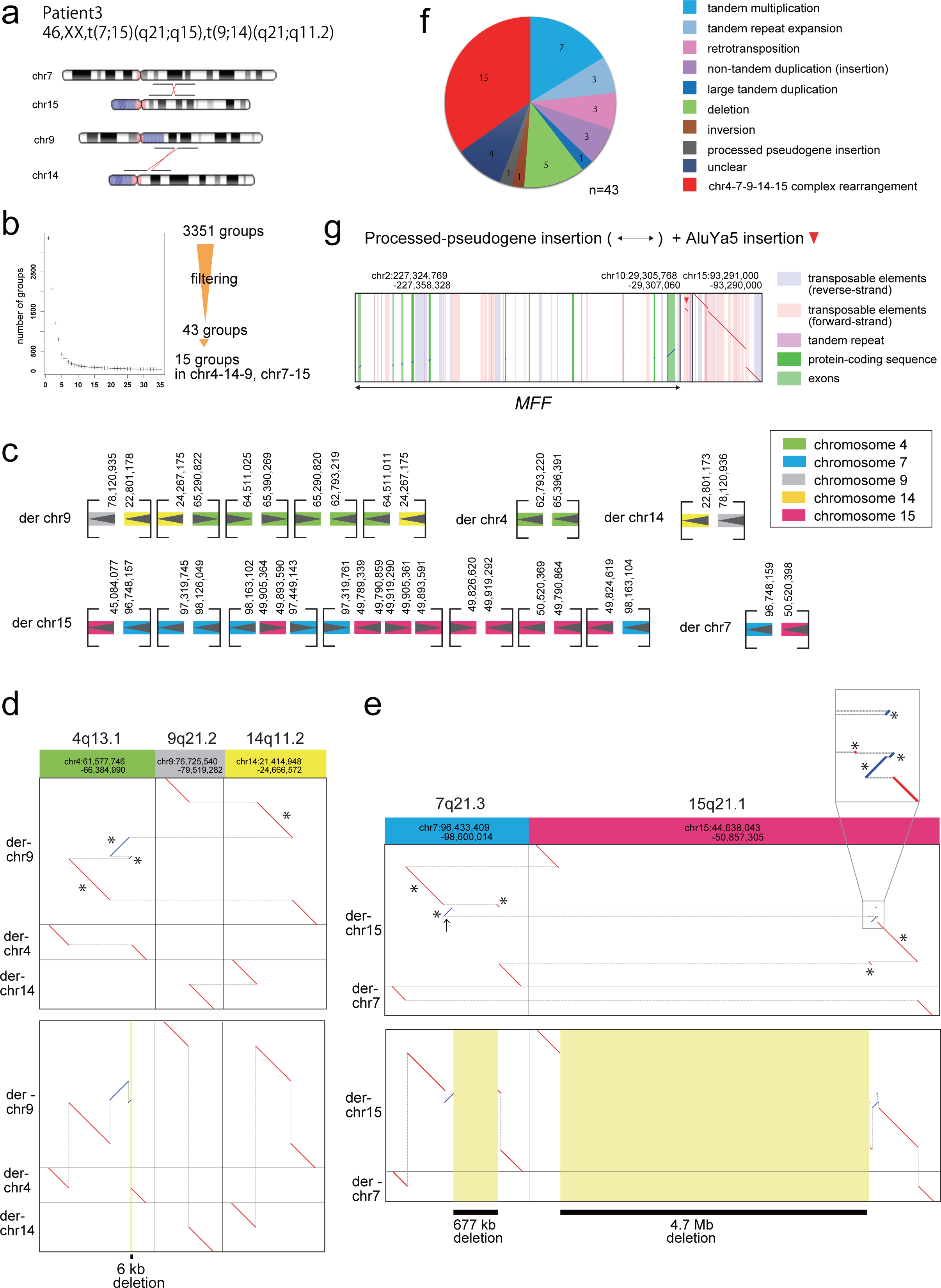
Chromosomal rearrangement in Patient 3 with 46,XX,t(7;15)(q21;q15),t(9;14)(q21;q11.2) a. Ideograms showing translocation positions of Patient 3. b. Filtering out rearrangements shared with 33 controls produces 43 groups of reads with patient-only rearrangements. While there are 2 groups indicating chr9-chr14 reciprocal translocation, there are 8 groups involved in the chr7-chr15 translocation. c. `dnarrange-link` with 5 additional groups involving chr14q11.2, 15 groups in total, which were linked to construct 5 derivative chromosomes. d. Dot-plot of reconstructed derivative chr9 and chr14 shows reciprocal balanced chromosomal translocation. Upper panel: horizontal dotted gray lines join the parts of each derivative chromosome. Asterisks indicate fragments. Lower panel: vertical dotted gray lines join fragments that come from adjacent parts of the reference genome, showing there is 6-kb deletion on chr4. e. Dot-plot of joined fragments showing reciprocal chr7-chr15 translocation with complex rearrangements. Black arrow indicates inverted 129 kb region that caused misinterpretation of deletion size. Upper panel: horizontal gray lines join the parts of each derivative chromosome. Asterisks indicate fragments. An inset magnifies 4 tiny fragments. Lower panel: vertical gray lines join fragments that come from adjacent parts of the reference genome, showing loss of 677 kb and 4.7 Mb of chr7 and chr15, respectively. f. Pie chart of patient-only rearrangements. g. Processed-pseudogene insertion in chr15 from exons of *MFF* on chr2, with nearby AluYa5 insertion.

**Figure 5.**
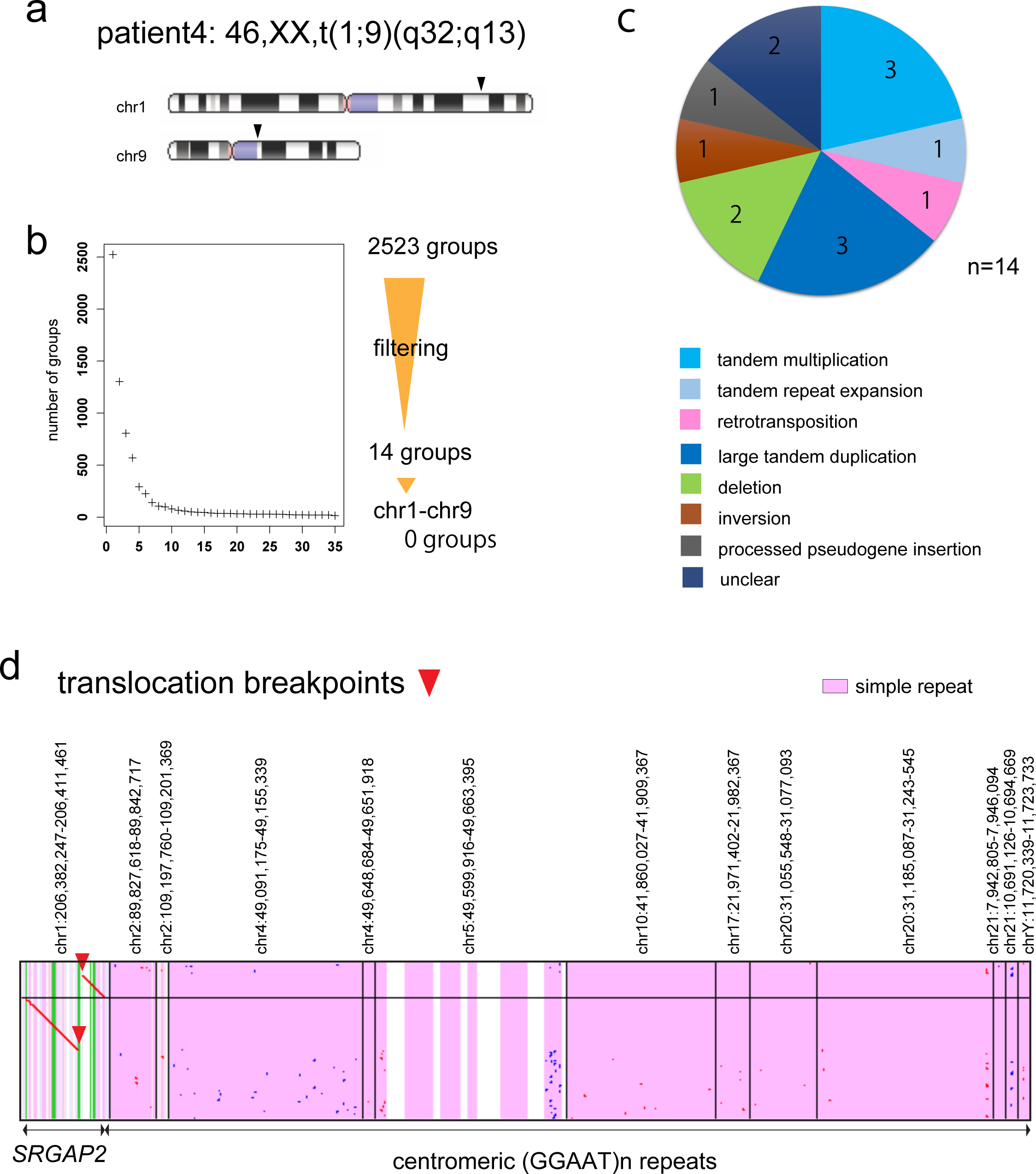
Chromosomal rearrangement in Patient 4 with 46,XX, t(1;9)(q32;q13) a. Ideograms showing translocation position of Patient 4. b. Filtering out rearrangements shared with controls produces 14 groups of reads with patient-only rearrangements. There is no group supporting chr1-chr9 translocation. c. Pie chart of patient-only rearrangements. d. Dot-plot of two reads that cross the chr1 breakpoint.

Computational time counts for this method (including filtering 33 controls) are shown in Supplementary Table 3. After filtering, we used `dnarrange-link` to infer the order and orientation of multiple groups, to understand the whole rearrangement (Fig2c, Fig3c, e, Fig4d, e).

### Patient 1

Patient 1 (Nishimura et al. described as Case2, and Bano et al. case report)^11,12^ has de novo reciprocal translocation between chr2 and chrX, 46,X,t(X;2)(q22;p13) (Fig2a). The breakpoints were not detected by short read sequencing^15^ though they were detected by more-painstaking breakpoint PCR^11^, so we tested whether we could find this rearrangement with long reads. We performed PromethION DNA sequencing (112 Gb), and found 2,773 groups of rearranged reads compared to human reference genome hg38. After subtracting rearrangements present in 33 controls, we found 80 patient-only groups, of which two involve both chr2 and chrX (Fig2b). These are exactly the reciprocal chr2-X translocation (Fig2c, Supplementary Fig 2). The breakpoints agreed with reported breakpoints determined by Sanger sequencing^11^.

The other 78 groups of rearranged reads are mostly: tandem multiplications (duplications, triplications, etc.), tandem repeat expansion/evolution, large deletions, retrotransposon insertions (five L1HS, four AluYa5, two AluYb8, three SVA, and one or two ERV-K LTRs), and other non-tandem duplications (Fig2d-e, Supplementary Table 4, 5, Supplementary Fig 3). These types of retrotransposon are known to be active or polymorphic in humans^16-18^. One case appears to be an orphan 3’-transduction from an L1HS in chr20: the L1HS was transcribed with readthrough into 3’ flanking sequence, then the 3’-end of this transcript (without any L1HS sequence) was reverse-transcribed and integrated into chr10 (Fig2e). Such orphan transductions can cause disease^19^. We also found an insertion of mitochondrial DNA (NUMT) into chr2 (Fig2e). Some of these rearrangements have been previously found in other humans, e.g. the ERV-K LTR inserted in chr12^20^. Thus our subtraction of rearrangements found in other humans was not thorough, especially because patient 1 is Caucasian whereas most of our controls (32/33) are Japanese.

### Patient 2

Patient 2 (Nishimura et al. described as Case1)^11^ has reciprocal chromosomal translocation between chr4 and chrX, 46,X,t(X;4)(q21.3;p15.2) and a 4 kb deletion of chrX and a 7 kb deletion of chr4 (Fig3a): these were found previously by Southern blot combined with inverse PCR sequencing^11^ but not by short read sequencing^15^. We performed PromethION DNA sequencing (117 Gb), and found 3,336 groups of rearranged reads relative to the reference genome, which reduced to 33 groups after control subtraction (Fig3b). Only 2 out of 33 groups involve both chr4 and chrX: they show a reciprocal unbalanced chromosomal translocation exactly as described previously ^11,15^ (Fig3c, Supplementary Fig 4). Another of the 33 groups shows a 43 kb deletion near the translocation site at chrX:107943791-107986323 (Fig3c, Supplementary Fig 4), which eliminates the *TEX13B* gene (Supplementary Fig 4), and was not previously described^15^. About half of the other rearrangements were tandem multiplication and retrotranspositions (Fig3d, Supplementary Fig 5, Supplementary Table 4, 5). Three of the 33 groups lie near each other in chr11q11 (Fig3e): they have a unique order and orientation that produces one linear sequence, whereby we fully inferred the structure of this previously-unknown rearrangement (Fig3e). The alignment dotplot (Fig3e) can be read from top to bottom, showing where each part of the rearranged sequence (vertical) comes from in the ancestral sequence (horizontal). This rearrangement has translocated and inverted fragments, and three deletions, including a 10kb deletion that removes most of the *TRIM48* gene.

### Patient 3: complex rearrangements at chr7-chr15 translocation

We next analyzed Patient 3 whose precise structure of chromosomal translocations was only partly solved before^13,15^. Patient 3 was reported to have two reciprocal chromosomal translocations between chr7 and chr15 as well as between chr9 and chr14, t(7;15)(q21;q15) and t(9,14)(q21;q11.2) (Fig4a), and has 4.6 Mb and ∼1 Mb deletions on chr15 and chr7, respectively, which were predicted by microarray, although the precise locations of breakpoints were not detected in detail. We performed whole genome nanopore sequencing (95 Gb) on this patient and found 3,351 groups of rearranged reads relative to the reference genome, which reduced to 43 groups after control subtraction (Fig4b). Fifteen out of 43 groups are involved in the two translocations: `dnarrange-link` found a unique way to order and orient them without changing the number of chromosomes (Fig4c, Supplementary Fig 6). At first, there seem to be two groups involving both chr9 and chr14, which accurately indicate the balanced chr9-chr14 translocation described previously^15^. However, `dnarrange-link` additionally identified a complex rearrangement for t(9,14)(q21;q11.2). A part of chr4 was unexpectedly inserted to derivative chr9 (Fig4d). This rearrangement was not investigated in the previous analyses, as chr7q21 was the primary locus for split-foot. In addition to this, `dnarrange` identified 8 out of 43 groups involving chr7 and chr15 (Fig4c, Supplementary Fig 6). The order and orientation of these groups was difficult to determine immediately by manual inspection, but `dnarrange-link` found only one possible way to connect them without changing the number of chromosomes (Fig4c). Finally, `dnarrange-link` could automatically reconstruct the whole rearrangements (Fig4d, e). The reconstructed rearrangements show that 3 fragments (breakpoint-to-breakpoint, asterisks in Fig4d, e) from chr4 and 1 fragment from chr14 were inserted into derivative chr9 (Fig4c, d), and 3 fragments from chr7 and 6 fragments from chr15 were inserted into derivative chr15 (Fig4c, e). They show 677 kb and 4.7 Mb deletions on chr7 and chr15, respectively, which were detected by microarray (Fig 4e). Note that these deletions are not present in any part of the rearrangement, but only in the fully-reconstructed rearrangement: they are holistic properties of the complex rearrangement. One candidate gene for split-foot, *SEM1* was not disrupted, nor had altered expression in lymphoblastoid cells (Supplementary Fig 7a, b, Supplementary Results).

A striking feature of these rearrangements is that the rearranged fragments come from near-exactly adjacent parts of the ancestral genome (Fig4d, e). This suggests that the rearrangements occurred by shattering of the ancestral genome into multiple fragments, which rejoined in a different order and orientation with loss of some fragments. Such shattering naturally explains why the fragments come from adjacent parts of the ancestor^8^.

We performed Sanger sequence confirmation for all 18 breakpoints (Supplementary Fig 8, primer sequences: Supplementary Table 6). There were only minor differences (usually 0 or 1 bases) between Sanger sequence-confirmed breakpoints and `dnarrange` predicted breakpoints from `lamassemble` consensus sequences (Supplementary Fig 9).

The other rearrangements are mostly local tandem-duplication or insertions (Supplementary Table 4, 5, Supplementary Fig 10). We found one processed pseudogene insertion, where exons of the *MFF* gene were inserted into chr15 (Fig4g). Interestingly, there is also an AluYa5 insertion into chr15 nearby (Fig4g). Both Alu and processed pseudogene insertions are thought to be catalyzed by LINE-1 encoded proteins: thus we speculate that these two insertions did not occur independently.

### Patient 4: difficult case with translocation breakpoint at centromere repeat

Patient 4 had a reciprocal translocation between chr1 and chr9 (Fig5a). Breakpoints in chr1 were previously described at chr1:206,401,153 and chr1:206,402,729, which disrupted *SRGAP2*, by intensive investigations using fluorescent in situ hybridization (FISH), Southern hybridization and inverse PCR^14^, or short read whole genome sequencing^15^. Chr9 breakpoints have not been found and were suspected to reside in repetitive centromeric heterochromatin. We performed PromethION DNA sequencing (41 Gb), and found 2,523 groups of rearranged reads relative to the reference genome, which reduced to 14 after control subtraction, none of which indicate chr1-chr9 translocation (Fig5b, c, Supplementary Table 5, Supplementary Fig 11). Dot-plot pictures of reads that cross the chr1 breakpoint suggest that there is a reciprocal translocation, but the other half of the read aligns (with low confidence) to satellite or simple repeat sequences at centromeric regions on multiple different chromosomes (Fig5d, two example reads are shown). This limitation might be overcome by obtaining reads long enough to extend beyond the centromeric repeats, or perhaps by obtaining a reference genome that is more accurate in centromeric regions.

## Discussion

We analyzed a variety of chromosomal translocations in 4 patients, who were selected because previous studies had difficulty in determining precise breakpoints by conventional approaches including microarrays and short read sequencing. Especially, complex rearrangements in Patient 3 were not solved even by intensive analysis^13,15^. Our method could not only precisely detect breakpoints but also characterize how shattered fragments were ordered and oriented. To the best of our knowledge, there has been no method to filter patient-only rearrangements, and connect them to reconstruct rearranged chromosomes by an automatic algorithm.

Recently, long read sequencing is becoming available for individual genome analysis due to a decrease in cost and increase in output data size. Accordingly, there have been a few approaches to use long read sequencing to detect structural variations^7,8,21^, including tandem-repeat changes in rare genetic diseases^6^, providing evidence that long read sequencing has a clear advantage in precisely detecting rearrangements. We observed that multiple breakpoints were jointly detected in a single read in Patient 3 (Supplementary Fig 8d, e), because long enough reads can cover several breakpoints, which is helpful to phase and order rearrangements. There are continuous efforts to obtain longer nanopore reads, however, in case of complex rearrangements (e.g. chromothripsis), it is not easy to cover whole rearrangements, as seen in Patient 3, by current long read sequencing read length. Our new tool, `dnarrange-link` is useful to infer a complete picture of complex rearrangements. In addition, `dnarrange-link` can provide a clear visualization of reciprocal chromosomal translocations, inversions or complex rearrangements with or without loss of sequences as seen in patients 1, 2 and 3. Most importantly, sequence loss was indicated after reconstructed derivative chromosomes were compared to the reference genome. We have shown that sequence losses in patient 3 agree with previously described microarray results. Previous studies on Patient 3 predicted 802 kb deletion (microarray could only suggest ∼1Mb deletion due to low resolution), because a small inversion (arrow in Fig4e) was missed by previous studies using long PCR. We also presented an example in Patient1, who has an inverted duplication on chr16, which was only understood as copy number gain, or simply inversion, by microarray or conventional sequencing technologies (Supplementary Fig 3, Supplementary Table 5). In summary, our approach using `dnarrange` and long read sequencing is superior to conventional approaches (e.g. microarray) because: it can 1) connect multiple rearrangements, 2) subtract shared rearrangements, and 3) detect balanced chromosomal rearrangements (e.g. inversion).

Our approach in this study narrowed down patient-only rearrangements using 33 controls. The number of rearrangements decreased exponentially with the first few samples to a few hundreds. This may be due to the presence of common rearrangements in the population. We suspect large numbers of controls will not be needed if there is a target rearrangement locus (e.g. 4p15.2) because the number of candidates is small. In all 4 patients, patient-only (not present in at least 66 autosomal alleles of 33 controls) rearrangements were fewer than 100. If we were to further narrow down to ultra-rare variations that may cause rare congenital disorders, a larger number of controls may be considered. Patient 1 has more patient-only groups of rearranged reads (80) than the other patients (33, 43 and 14). This is because the patient is Caucasian and most of the control data used were Japanese (32/33 datasets). Applying ethnicity-matched controls, or parents or other relatives, will be useful to further remove benign rearrangements.

We noticed that large fractions of these rearrangements are insertions or tandem multiplications (Supplementary Table 4). Perhaps surprisingly, patient-specific simple inversions were uncommon. There are several types of insertions which are also known to cause diversity of human genomes^22^ e.g. transposable element (TE) insertions, especially L1HS, AluYa5 or AluYb8^23^, ERV^24^, nuclear mitochondrial DNA insertions (NUMT)^25^, or processed pseudogene insertions^26^ (Supplementary Results, Supplementary Fig 12, Supplementary Table 7). Interestingly, most of the inserted sequences were aligned to TEs. TE insertions may be a common type of rare variation seen in individuals. In addition to TE-insertion, we detected rare processed pseudogene insertion in 3 patients. Two of these insertions were previously described with allele frequency 1-10% in Japanese (*MFF*) and 1-10% in non-Japanese (*MATR3*)^26^. We also observed non-tandem duplications that do not seem to be retrotranspositions: interestingly, about half of these are localized, i.e. a copy of a DNA segment is inserted near (e.g., within a few kb of) the original segment^8^ (see blue highlighted loci in Supplementary Table 5).

Our analysis proves useful despite its dubious assumption that the reference genome is ancestral to the DNA reads. This may be partly because we focus on disease-causing rearrangements, which are likely to be derived. Also, incorrect rearrangements due to a non-ancestral reference may be found in both patients and controls, thus filtered out. It would be useful to construct a reference human genome that is ancestral (and complete), as far as possible, because this simplifies the relationship between the reference and extant human DNA sequences^8^.

Our method in combination with subtracting shared rearrangements in control datasets has a great strength in precisely detecting chromosomal rearrangements, including inversions, translocations, TE insertions, NUMT and processed pseudogene insertions. There has been no method that can effectively subtract rearrangements shared in the population, thus we believe our method is useful to analyze complex rearrangements in a clinical setting (i.e. rare genetic disease or perhaps cancer genomes). We also showed a limitation of our method: detecting rearrangements in large repetitive regions beyond the length of long reads in Patient 4. These regions are still elusive and highly variable between individuals. To date there is no good method to detect rearrangements in large repetitive regions (e.g. centromeric or telomeric repeats) genome-wide. We hope our understanding of these still-intractable regions will expand as sequencing technologies advance.

In conclusion, we developed an effective method to find chromosomal aberration, with precise breakpoint identification, only from long read sequencing. Our method also provides an automatic algorithm for reconstruction of complex rearrangements. Long read sequencing may be considered when chromosomal abnormalities are suspected.

## Methods

### Samples and ethical issues

All genomic DNA from patients and controls were examined after obtaining informed consent. Experimental protocols were approved by institutional review board of Yokohama City University under the number of A19080001.

#### dnarrange

`dnarrange` finds DNA reads that have rearrangements relative to a reference genome, and discards “case” reads that share rearrangements with “control” reads (Supplementary Methods). It takes one or more files of read-to-genome alignments, where each file is a “case” or a “control”. It assumes the alignments have this property, which is guaranteed by `last-split`: each read base is aligned to at most one genome base. `dnarrange` first performs these steps, for cases and controls:

1. In order to recognize large “deletions” as rearrangements, if an alignment has deletions >= *g* (a threshold; default 10kb), split it into separate alignments either side of these deletions.
2. Get rearranged reads. We classify rearrangements into four types: inter-chromosome, inter-strand (if a read’s alignment jumps between the two strand of a chromosome), non-colinear (if a read’s alignment jumps backwards on the chromosome), and “big gap” (if a read’s alignment jumps forwards on the chromosome by >= g).
3. Discard any “case” read that shares a rearrangement with any “control” read. (Two reads are deemed to share a rearrangement if they have similar rearrangements that overlap in the genome: the precise criteria are in the Supplementary methods, Supplementary Fig 13, 14.) It then performs these steps, for cases only:
4. Discard any read with any rearrangement not shared by any other read. Repeat this step until no further reads are discarded (so that `dnarrange` has the useful property of *idempotence*).
5. Group reads that share rearrangements. First, a link is made between any pair of reads that share a rearrangement. Then, groups are connected components, i.e. sets of reads linked directly or indirectly.
6. Discard groups with fewer than 3 reads.

#### dnarrange-link

`dnarrange-link` infers how the rearranged fragments found by `dnarrange` are linked to each other, and thereby reconstructs the derived chromosomes. It uses (the alignments of) one representative read per group. The representative could be one actual read, or a consensus sequence. Based on the alignments, the two ends of each read are classified as “left” if the alignment extends rightwards/downstream along the chromosome starting from that end (shown as “[“in Fig6a) or “right” if the alignment extends leftwards/upstream (“]”). Two ends may be directly linked only if:

**Figure 6.**
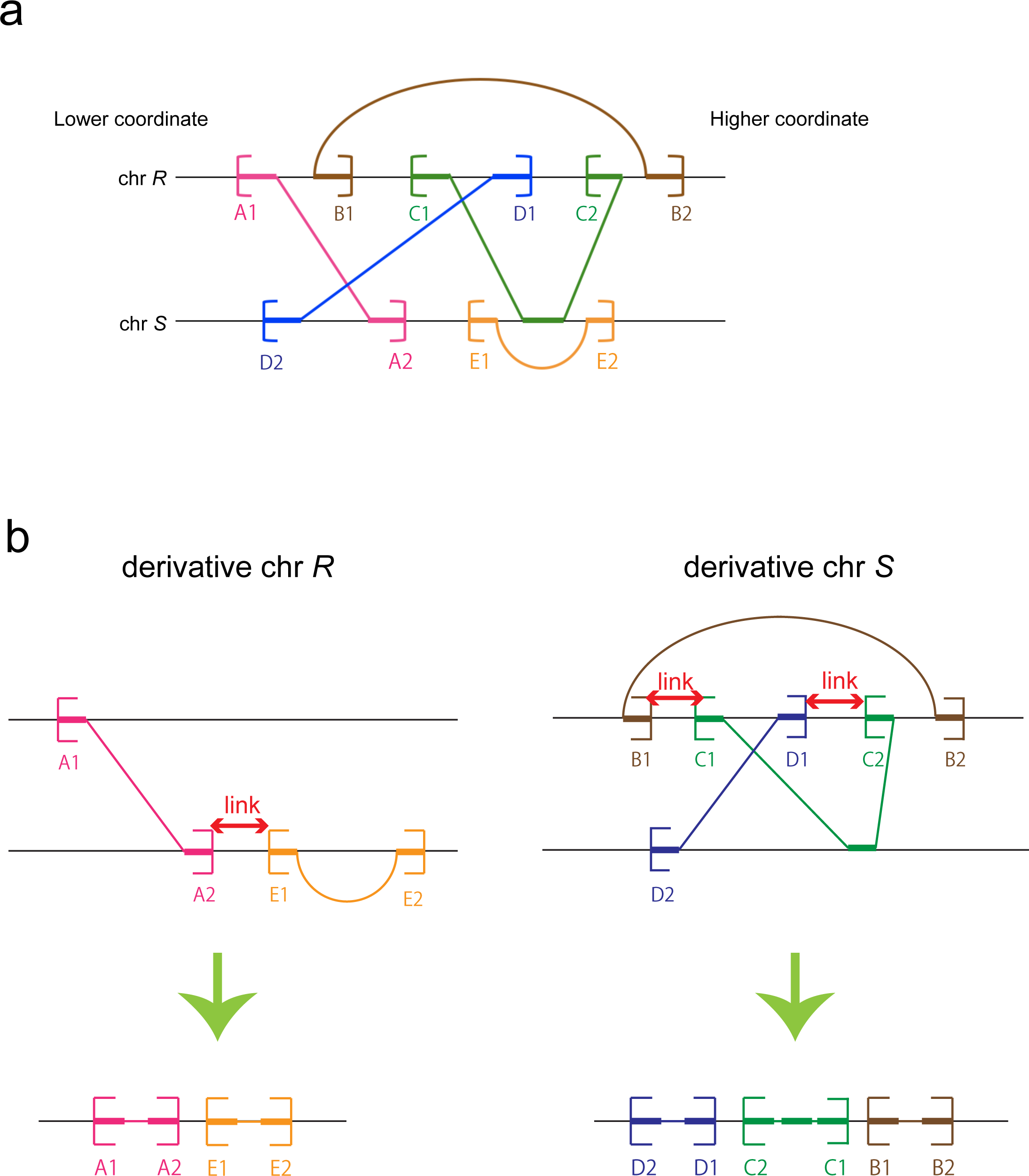
Illustration of data analyzed by dnarrange-link. a. The sketch shows alignment of five DNA reads (A, B, C, D, E) to a genome. The two ends of each read are arbitrarily labeled 1 and 2. b. Derivative chr *R* was reconstructed by linking A2 to E1 (left). Derivative chr *S* was reconstructed by linking B1 to C1, and D1 to C2 (right). B1 can also be linked to C2, but in that case it is impossible to link C1 to anything, and D1 to anything, thus this possibility was suppressed.

^*^ They are on the same reference chromosome.

^*^ One is left and the other is right.

^*^ The left end is downstream of (has higher reference coordinate than) the right end.

In order to infer the actual links, we require some further information or assumption. We make this assumption: there are as many links as possible, or equivalently, the derived genome has as few chromosomes as possible. For example, in Fig6a, B1 may be linked to C2, but in that case it becomes impossible to link C1 to anything, and D1 to anything. Based on our assumption, we instead link B1 to C1 and D1 to C2. In this example, `dnarrange-link` infers two derivative chromosomes: one is reconstructed from two reads by linking A2 to E1, the other is reconstructed from three reads by linking D1 to C2 and C1 to B1 (Fig6b).

The two types of end, with linkability relationship, define a bipartite graph. To infer the links based on our assumption, we find a “maximum matching” in this graph. If there is more than one maximum matching, one is chosen arbitrarily, and a warning message is printed. In Fig6, there is only one maximum matching.

In Fig6a, the left and right ends occur in an alternating pattern along each reference chromosome. In this case, we get a unique maximum matching by linking adjacent left and right ends. This alternating pattern seems to occur often in practice.

#### lamassemble

`lamassemble` merges overlapping DNA reads into a consensus sequence, by these steps (details in the Supplement):

1. Calculate the rates of insertion, deletion, and substitutions between two reads by “doubling” the rates from last-train, because errors occur in both reads.
2. Use these rates to find pairwise alignments between the reads with LAST. LAST also calculates the probability that each pair of bases is wrongly aligned (which is high when there are alternative alignments with near-equal likelihood).
3. Use the LAST alignments in descending order of score to define a tree for progressive alignment by MAFFT.
4. Constrain the MAFFT alignment by anchoring pairs of bases that were aligned by LAST with error probability <= 0.002.
5. Make a consensus sequence from the MAFFT alignment. Omit alignment columns with gaps in > 50% of sequences covering that column. For each column, get the base that maximizes prob(base|column), using the last-train substitution probabilities.

Some results using a prototype of `lamassemble` were published previously^6^.

### Nanopore sequencing using PromethION

DNA was extracted from patients’ blood cells. Libraries were prepared for nanopore sequencing using DNA ligation kit (SQK-LSK109) then subjected to PromethION sequencing (Oxford Nanopore Techonologies) using one PRO-002 (R9.4.1) flowcell according the manufacturer’s protocol. Base-calling and fastq conversion were performed with MinKNOW ver1.14.2. Control datasets were also sequenced by PromethION. Base-calling and fastq conversion were performed with MinKNOW ver1.11.5.

### Sanger-sequence confirmation of breakpoints

PCR primers for breakpoints estimated from rearrangements were designed using primer3 plus software (Supplementary Table 6). PCR amplification was done using ExTaq (Takara) and LATaq, then amplified products were Sanger sequenced using BioDye Terminator v3.1 Cycle Sequencing kit with 3130xl genetic analyzer (Applied Biosystems, CA, USA).

## Data Availability

All data are private and not available.

## Web resources

LAST: http://last.cbrc.jp

MAFFT: https://mafft.cbrc.jp/alignment/software/

lamassemble: https://gitlab.com/mcfrith/lamassemble

dnarrange: https://github.com/mcfrith/dnarrange

NCBI genome decoration: https://www.ncbi.nlm.nih.gov/genome/tools/gdp

Primer3: http://bioinfo.ut.ee/primer3-0.4.0/

UCSC genome browser: https://genome.ucsc.edu/

## Acknowledgements

This work was supported by AMED under the grant numbers JP19ek0109280, JP19dm0107090, JP19ek0109301, JP19ek0109348, JP18kk020501 (to N. Matsumoto) and JP19am0101108 (to K. Katoh); JSPS KAKENHI under the grant numbers JP17H01539 (to N. Matsumoto), JP16K07464 (to K. Katoh) and JP19K07977 (to S. Mitsuhashi); grants from the Ministry of Health, Labor, and Welfare (to N. Matsumoto); and the Takeda Science Foundation (to N. Matsumoto).

## Author contributions

OS, SM, MCF, KK and NM contributed to the conception of the work and acquisition/analysis/interpretation of the data.

